# A Distinct Inflammatory Milieu in Patients with Right Heart Failure

**DOI:** 10.1101/2023.04.12.23288502

**Authors:** Bin Q Yang, Arick C Park, Jason Liu, Kathleen Byrnes, Ali Javaheri, Douglas L Mann, Joel D Schilling

## Abstract

**Background:** Right heart failure (RHF) is associated with worse clinical outcomes. In addition to hemodynamic perturbations, the syndrome of RHF involves liver congestion and dysfunction. The mechanisms that underlie heart-liver interactions are poorly understood and may involve secreted factors. As a first step to understand the cardiohepatic axis, we sought to elucidate the circulating inflammatory milieu in patients with RHF.

**Methods:** Blood samples were collected from the IVC and hepatic veins during right heart catheterization from 3 groups of patients: 1) controls with normal cardiac function, 2) patients with heart failure (HF) who did not meet all criteria of RHF, and 3) patients who met prespecified criteria for RHF defined by hemodynamic and echocardiographic parameters. We performed multiplex protein assay to survey levels of several circulating markers and analyzed their association with mortality and need for left ventricular assist device or heart transplant. Finally, we leveraged publicly available single cell RNA sequencing (scRNAseq) data and performed tissue imaging to evaluate expression of these factors in the liver.

**Results:** In this study of 43 patients, RHF was associated with elevated levels of a subset of cytokines/chemokines/growth factors compared to controls. In particular, soluble CD163 (sCD163) and CXCL12 were higher in RHF and predicted survival in an independent validation cohort. Furthermore, scRNAseq and immunohistochemistry of human liver biopsies suggest that these factors are expressed by Kupffer cells and may be liver derived.

**Conclusions:** RHF is associated with a distinct circulating inflammatory profile. sCD163 and CXCL12 are novel biomarkers that can prognosticate patient outcomes. Future studies to define how these molecules influence HF phenotypes and disease progression may lead to new approaches to management of patients with RHF.

## What is new?

• Right heart failure (RHF) is a strong predictor of poor clinical outcomes, although the mechanisms are not well understood. We identified a unique set of circulating inflammatory markers in patients with RHF that are associated with increased risk of adverse events.

## What are the clinical implications?

• We provide evidence that liver resident macrophages are altered in RHF and that they serve as a source inflammatory factors found in patients with RHF. Biomarkers associated with the cardiohepatic axis may help inform decision-making between patients and physicians and drive new discoveries to improve clinical outcomes in patients with heart failure.

## Non-standard Abbreviations and Acronyms

Right heart failure – RHF Heart failure – HF

Central venous pressure – CVP Congestive hepatopathy – CH

Left ventricular assist device – LVAD Heart transplant – HT

Kupffer cells – KC

Single cell RNA sequencing – scRNAseq Right heart catheterization – RHC

Right atrial pressure – RAP

Pulmonary capillary wedge pressure – PCWP Tricuspid annular plane systolic excursion – TAPSE Left ventricular – LV

Ejection fraction – EF

Receiver-operating characteristic – ROC Area under curve – AUC

Pulmonary vascular resistance – PVR Vascular endothelial growth factor – VEGF Soluble CD163 – sCD163

Right ventricular – RV Stromal-derived factor – SDF

## INTRODUCTION

Right heart failure (RHF) is associated with morbidity and mortality in patients with heart failure (HF) and pulmonary hypertension.^1, 2^ However, the specific mechanisms that underlie this relationship are less clear. In patients with RHF, persistently elevated central venous pressure (CVP) contributes to congestive hepatopathy (CH), cardiorenal syndrome, and increased intestinal permeability.^3–5^ Furthermore, RHF is associated with increased risk of complications such as gastrointestinal bleeding after left ventricular assist device (LVAD) implantation, primary graft dysfunction after heart transplant (HT), and vasoplegia after advanced HF surgeries.^6, 7^ These observations raise the possibility that circulating factors that modulate inflammation and angiogenesis may be altered in RHF. Although HF has been associated with elevated proinflammatory cytokines, the specific influence of RHF on inflammation has not been previously investigated.^8, 9^

The liver sits at the intersection between the systemic and splanchnic circulation and plays an important role in homeostasis and immune regulation.^10, 11^ Advanced liver disease has been associated with increased mortality in patients with HF and is a relative contraindication to LVAD or HT.^12, 13^ Yet, the cause of death in these patients is rarely liver failure, suggesting that systemic consequences of CH, such as dysregulated immune activation, may be contributing to this phenomenon. In the setting of RHF, the liver is exposed to both hemodynamic stress from high CVP and inflammatory stress from translocation of gut microbial antigens, both of which could alter liver inflammatory responses. Moreover, the liver contains the largest reservoir of macrophages, known as Kupffer cells (KC), in the body. KC are located intravascularly within the hepatic sinusoids, where they are uniquely positioned to respond to both mechanical and microbial stress.^4^ However, the key pathways, molecular effectors, and biomarkers in RHF and CH are not well characterized.

To gain insight into the inflammatory signature in the syndrome of RHF, we measured circulating cytokines and growth factors in patients with RHF and compared them to controls and those with predominant left-sided HF. We uncovered a panel of inflammatory molecules that were elevated in HF and RHF, of which two had independent prognostic value in separate HF populations. To complement this analysis, we utilized single cell RNA sequencing (scRNAseq) data and immunohistochemistry of liver biopsy tissue from patients with CH to provide evidence that some of these factors may represent liver stress and KC activation.

## METHODS

### Data disclosure statement

The data that supports the findings of this study are available from the corresponding author upon reasonable request.

### Patient selection and follow up

Adult patients between ages 18-75 referred for right heart catheterization (RHC) at Washington University School of Medicine in St. Louis were screened. Patients with LVAD or HT, autoimmune conditions, active infection, cancer in remission for less than 5 years, or known chronic liver disease from another etiology were excluded. Informed consent was obtained prior to their procedure. All patients were followed for at least 6 months after RHC and the last date of censor was 3/1/2022. This study was approved by the Washington University Institutional Review Board (IRB# 201903133).

### RHF definition

Patients who consented for this study were classified into normal, HF, or RHF cohorts using echocardiographic and hemodynamic criteria that were defined *a priori.* Normal controls did not have any evidence of cardiac disease. Inclusion in the RHF cohort required all three of the following metrics: right atrial pressure (RAP) > 9mmHg, RAP/pulmonary capillary wedge pressure (PCWP) ratio > 0.55, and tricuspid annular plane systolic excursion (TAPSE) < 1.9cm.^14, 15^ HF patients had clinical HF but did not meet all three of the aforementioned criteria.

### RHC and blood sampling

Patients underwent RHC per routine clinical practice. Briefly, the right internal jugular or femoral vein was accessed using a 7Fr or 8Fr sheath. A 7Fr TD or 7.5Fr VIP Swan-Ganz catheter (Edwards LifeSciences) was inserted through the sheath and used to sample 10mL blood in the IVC below the hepatic veins. Subsequently, the catheter was advanced to the hepatic vein wedge position under fluoroscopic guidance per balloon-tip technique as previously described and an additional 10mL blood was collected.^16^ For technically difficult cases, a 0.025in diameter, 260cm Swan-Ganz catheter wire (Cook Medical) was employed at the discretion of the attending physician. After simultaneous blood sampling from the IVC and hepatic vein, the rest of the procedure was conducted per standard protocol.

### Inflammatory biomarkers

Samples were immediately centrifuged at 2000 RPM for 15 minutes to pellet the red blood cells and serum were aliquoted and stored in −80C until analysis. Measurement of inflammatory biomarkers was performed using Luminex Flexmap 3D platform housed within the Immunomonitoring Core Lab at Washington University. All samples were analyzed at the same time to prevent batch effects and ran in duplicates with 7-point internal quality control. Commercial assays were purchased from ThermoFisher (Human 45-plex panel, EPXR450-12171-901) and R&D Systems (Human Luminex discovery assay, LXSAHM-01).

### Validation cohort

To validate our findings in an external cohort, we utilized samples from the Washington University HF Registry. Briefly, this is a prospective registry of ambulatory HF patients across a spectrum of left ventricular (LV) ejection fraction (EF) and 172 patients within this registry underwent SOMAscan proteomics on peripheral blood samples. The methods and results from this study have been previously published.^17^ We retrospectively analyzed the association between elevated inflammatory biomarkers and mortality and HT-free survival.

### scRNAseq

Liver scRNAseq data was obtained from the publicly available human liver cell atlas (https://www.livercellatlas.org). Briefly, liver tissue was obtained in patients undergoing liver resection, cholecystectomy, or bariatric surgery and CITE-seq was performed on single cell and single nuclear preparations.^18^

### Liver biopsy immunohistochemistry/immunofluorescence

We queried liver biopsies performed between 2019-2021 with “congestive hepatopathy” in the pathology report and confirmed clinical HF diagnosis by chart review. We stained for macrophage markers CD68 (Ventana Medical Systems, 790-2931) and CD163 (Cell Marque, 163M-1) on 10 HF and 5 control samples in collaboration with the Anatomic and Molecular Pathology Core Lab. Quantification of CD163 area and cell count was performed using ImageJ software (ImageJ, National Institute of Health) and reported as an average over four random 40x high powered fields per sample. For immunofluorescence, paraffin-embedded liver tissue from HF patients were stained with rat anti-human CD68 (BioRad, MCA1957; 1:250 dilution) and mouse anti-human CXCL12 (R&D Systems, MAB350; 1:50 dilution) primary antibodies. Secondary antibodies were anti-mouse Alexa480 and anti-rat Alexa594 (Jackson Labs, 1:500 dilution) and nuclei were stained with Hoechst dye (1:25,000) for 5 minutes in the dark. The sections were then mounted with ProLong Gold Antifade reagent and a coverslip. Confocal images were taken using an LSM 700 laser scanning confocal microscope (ZEISS; Jena, Germany) with 10x 0.3 N.A. or 20x 0.8 N.A. objective at ambient temperature using the same laser intensity and gain with each magnification.

### Statistical analysis

Categorical and continuous variables were compared using Fisher’s exact and Student’s t-tests, respectively. For comparison between more than 2 groups, one-way ANOVA was performed. Shapiro-Wilk test was utilized to assess for normality and non-normal distributions were log-transformed. Due to the exploratory nature of our study, we generated Kaplan-Meier curves for mortality, stratified by the median value in the RHC cohort, and compared them using the log-rank test. For multivariable analysis, we utilized a Cox proportional hazards model adjusted for age, creatinine, NT-proBNP, and EF in the RHC cohort and age, gender, race, creatinine, and EF in the validation cohort, due to lack of NT-proBNP data and larger sample size in the latter. All statistical analyses were performed using GraphPad Prism 9.3.0 (GraphPad Software, San Diego, CA).

## RESULTS

### Baseline characteristics

We included 43 patients in this study and median time of follow up was 321 days. Compared to normal controls (n=9), patients with HF (n=17) and patients with RHF (n=17) had higher NT-proBNP, bilirubin, creatinine, and lower EF and cardiac index **(Table 1)**. The RHF cohort had even higher RAP and RAP/PCWP ratio. The majority of patients underwent RHC in the inpatient setting for advanced HF therapy evaluation and/or to assist with clinical management. Seven patients died (16%) and 16 patients received an LVAD/HT (37%) during follow up. LVAD/HT-free survival did not differ when stratified by EF or pulmonary vascular resistance (PVR) in patients with HF or RHF **(Figure 1)**.

**Figure 1.**
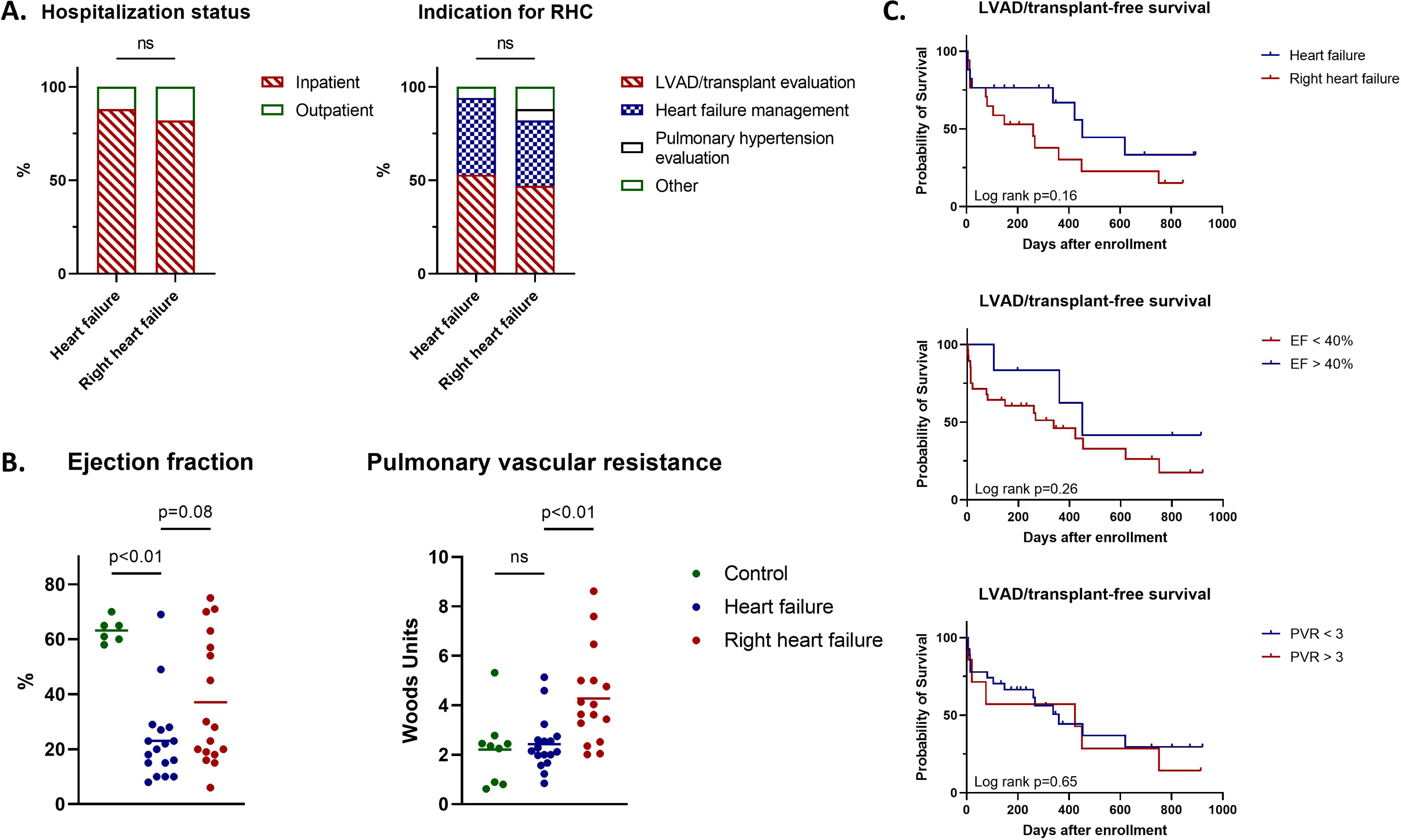
Comparison of heart failure (HF) and right heart failure (RHF) patients. **(A)** Distribution of patients based on hospitalization status and indication for right heart catheterization (RHC). **(B)** Distribution of left ventricular ejection fraction (EF) and pulmonary vascular resistance (PVR) across the control (green), HF (blue), and RHF (red) cohorts. The bars represent mean values. **(C)** Kaplan-Meier survival analysis of patients with HF based on presence of RHF (top), LVEF > 40% (middle), or PVR > 3 Wood Units (bottom). Log rank p-values are shown.

**Table 1.** Baseline clinical characteristics of normal, heart failure (HF), and right heart failure (RHF) patients. Categorical variables are expressed as n(%) and continuous variables are expressed as mean(SD). RAP=right atrial pressure, TAPSE=tricuspid annular plane systolic excursion. *p<0.05 between normal and HF and ^†^p<0.05 between HF and RHF cohorts.

|  | Normal (n=9) | HF (n=17) | RHF (n=17) |
| --- | --- | --- | --- |
| Age (years) | 49 (17) | 53 (7) | 57 (12) |
| Sex |  |  |  |
| Male | 3 (33) | 13 (76) | 9 (53) |
| Female | 6 (66) | 4 (24) | 8 (47) |
| Race |  |  |  |
| White/Caucasian | 7 (78) | 9 (53) | 9 (53) |
| Black/African-American | 2 (22) | 8 (47) | 8 (47) |
| HF etiology | -- |  |  |
| Ischemic |  | 3 (18) | 4 (24) |
| Non-ischemic |  | 14 (82) | 13 (76) |
| NT-proBNP (pg/mL) | 188 (58) | 2830 (2843) | 2927 (2093) |
| Creatinine (mg/dL) <sup>*†</sup> | 0.8 (0.2) | 1.2 (0.3) | 1.7 (0.7) |
| Bilirubin (mg/dL) | 0.4 (0.3) | 1.1 (1.0) | 1.3 (1.1) |
| RAP (mmHg) <sup>*†</sup> | 6 (3) | 11 (4) | 19 (5) |
| RAP/Pulmonary capillary wedge pressure <sup>†</sup> | 0.52 (0.16) | 0.52 (0.15) | 0.79 (0.18) |
| Pulmonary vascular resistance (Wood Units) <sup>†</sup> | 2.2 (1.4) | 2.4 (1.1) | 4.3 (1.9) |
| Cardiac index (L/min/m<sup>2</sup>) <sup>*</sup> | 3.4 (1.4) | 2.2 (0.5) | 2.0 (0.6) |
| Left ventricular ejection fraction (%) <sup>*</sup> | 63 (4) | 23 (15) | 37 (23) |
| Left ventricular end diastolic diameter (cm) <sup>*†</sup> | 4.5 (0.6) | 6.9 (1.1) | 5.8 (1.5) |
| Right ventricular end diastolic diameter (cm) | 3.9 (0.7) | 4.0 (1.1) | 4.6 (0.9) |
| TAPSE (cm) <sup>*</sup> | 2.5 (0.3) | 1.7 (0.6) | 1.4 (0.3) |
| Hospitalization status (%inpatient) | -- | 15 (88) | 14 (82) |
| Indication for RHC |  |  |  |
| Advanced HF evaluation | -- | 9 (53) | 8 (47) |
| HF management | -- | 7 (41) | 6 (35) |
| Pulmonary hypertension evaluation | 2 (22) | 0 (0) | 1 (6) |
| Other | 7 (78) | 1 (6) | 2 (12) |

### RHF is associated with increased circulating inflammatory cytokines and growth factors

To evaluate the circulating inflammatory milieu in patients with HF and RHF, we analyzed serum samples collected at time of RHC using a multiplex protein assay that contained a broad representation of cytokines, chemokines, and growth factors. We observed a distinct secretome in patients with HF and RHF **(Figures 2,3, Supplemental Figure 1).** Compared to controls, patients with RHF had elevated levels of a number of inflammatory cytokines and chemokines including TNFα (p=0.03), MIP-1β (p<0.01), CXCL10 (p<0.01), and CXCL12 (p<0.01). Moreover, RHF patients had higher levels of several growth factors including vascular endothelial growth factor-A (VEGF-A, p<0.01), hepatocyte growth factor (HGF, p<0.01), and stem cell factor (SCF, p=0.01). The levels of soluble CD163 (sCD163), IL-7, IL-8, and IL-18 were also increased in RHF with borderline p values (p=0.06-0.10). Although several markers showed trends for increased levels in patients RHF compared to HF, only CXCL12 was significantly different between these groups (p<0.01). There was no measurable difference in biomarkers between samples collected in the IVC versus hepatic veins.

**Figure 2.**
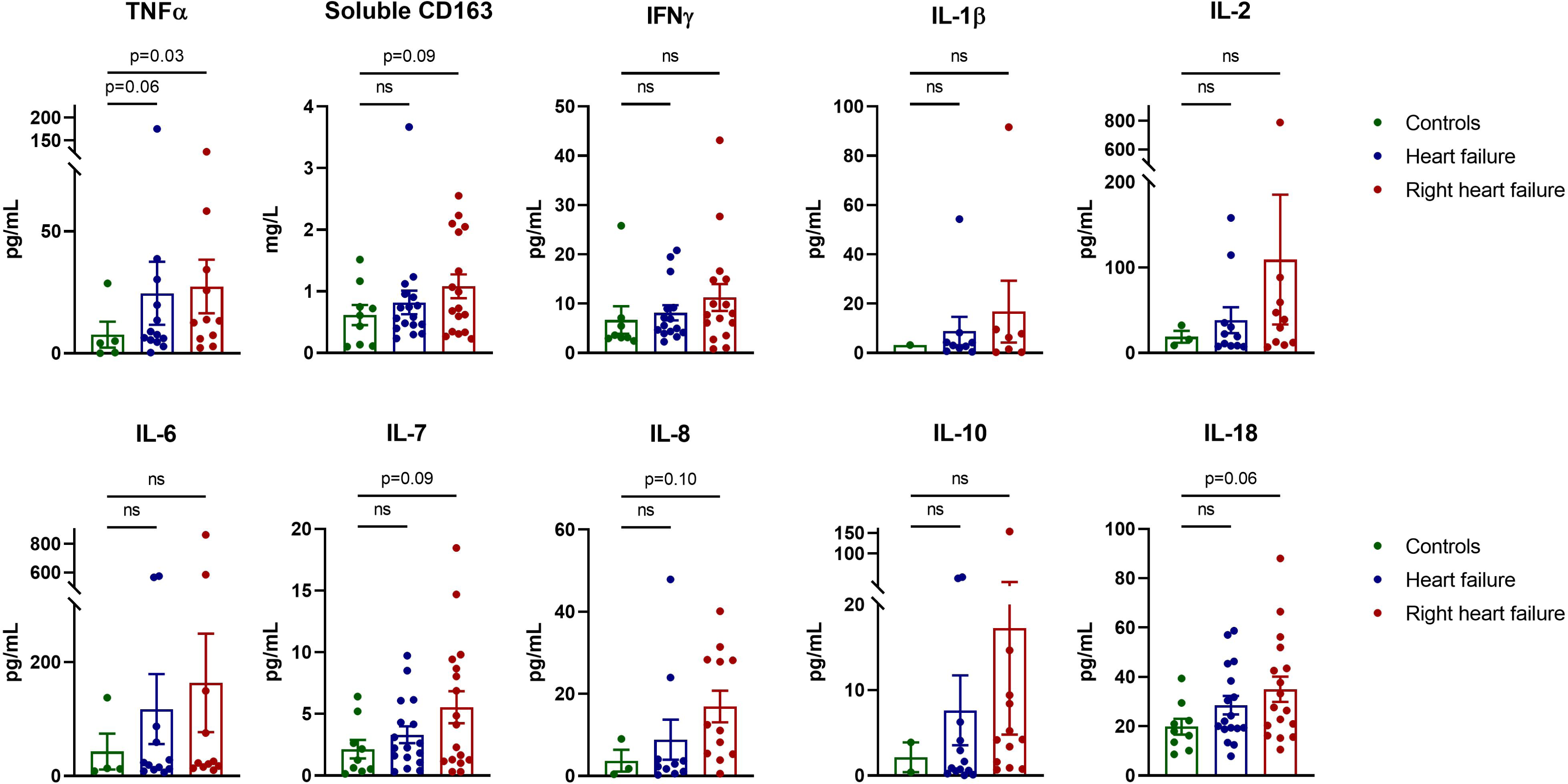
Cytokine profile in control, heart failure (HF) and right heart failure (RHF) patients. Quantification of the indicated cytokines via multiplex protein assay in control (green), HF (blue), and RHF (red) cohorts. The dots represent individual patient values, with the mean and standard deviation shown. P-values by ANOVA are shown for the indicated comparisons.

**Figure 3.**
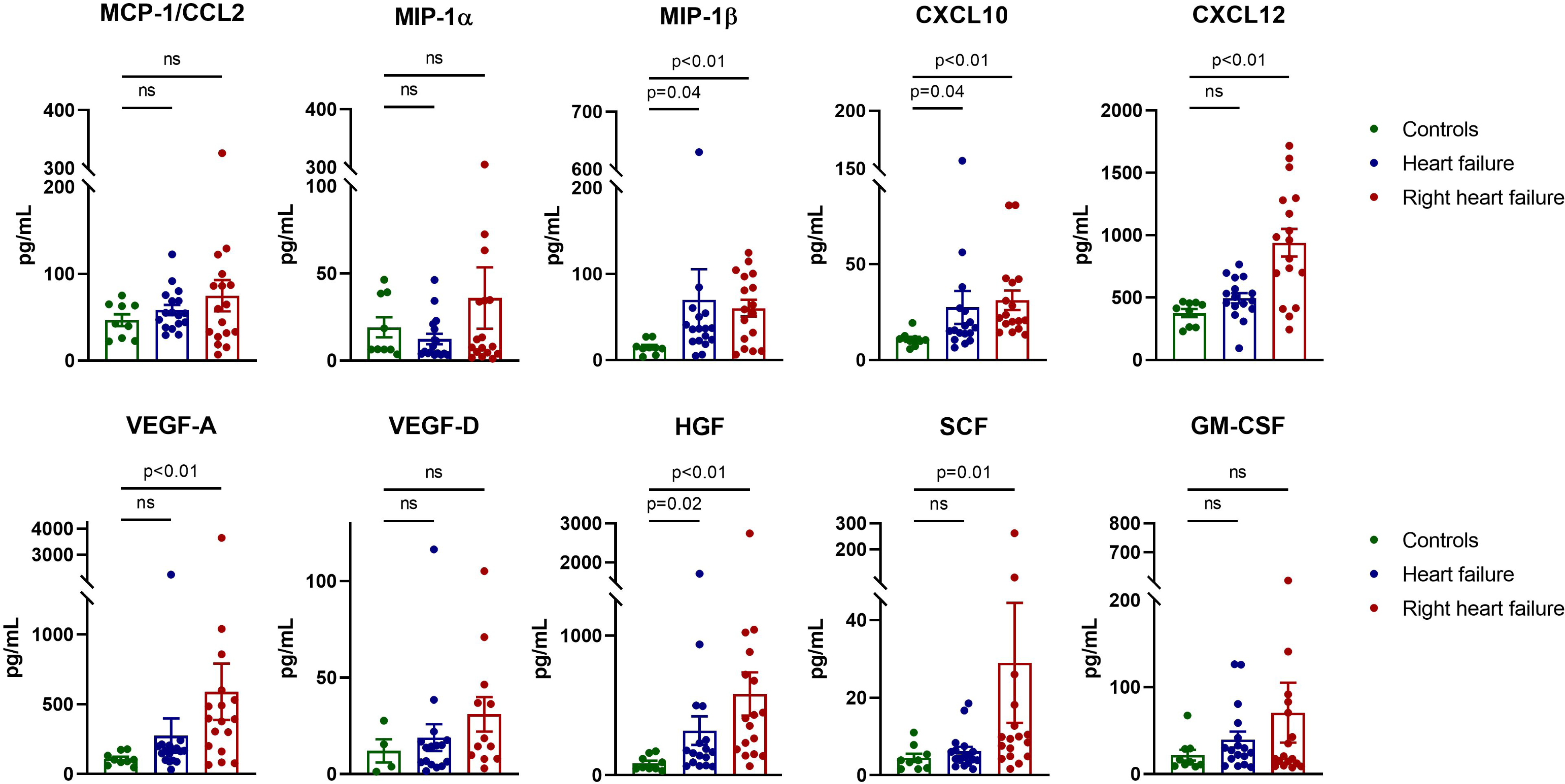
Chemokine and growth factor profile in control, heart failure (HF) and right heart failure (RHF) patients. Quantification of the indicated chemokines and growth factors via multiple protein assay in control (green), HF (blue), and RHF (red) cohorts. The dots represent individual patient values, with the mean and standard deviation shown. P-values by ANOVA are shown for the indicated comparisons.

### VEGF-A, sCD163, and CXCL12 predicted clinical outcomes

We next sought to determine the prognostic value of the serum inflammatory cytokines and growth factors that were elevated or had a strong trend towards higher levels in patients with RHF. We generated Kaplan-Meier curves for all-cause mortality stratified by the median value of the biomarker and performed Cox proportional hazards analysis adjusted for age, creatinine, NT-proBNP, and EF. We found that higher VEGF-A (HR 2.52, CI 0.80-5.87) and sCD163 (HR 1.83, CI 0.86-3.37) trended towards significance, and CXCL12 (HR 17.1,CI 3.97-84.3) was associated with increased hazard of death, LVAD, and HT **(Figure 4, Supplemental Figure 2).**

**Figure 4.**
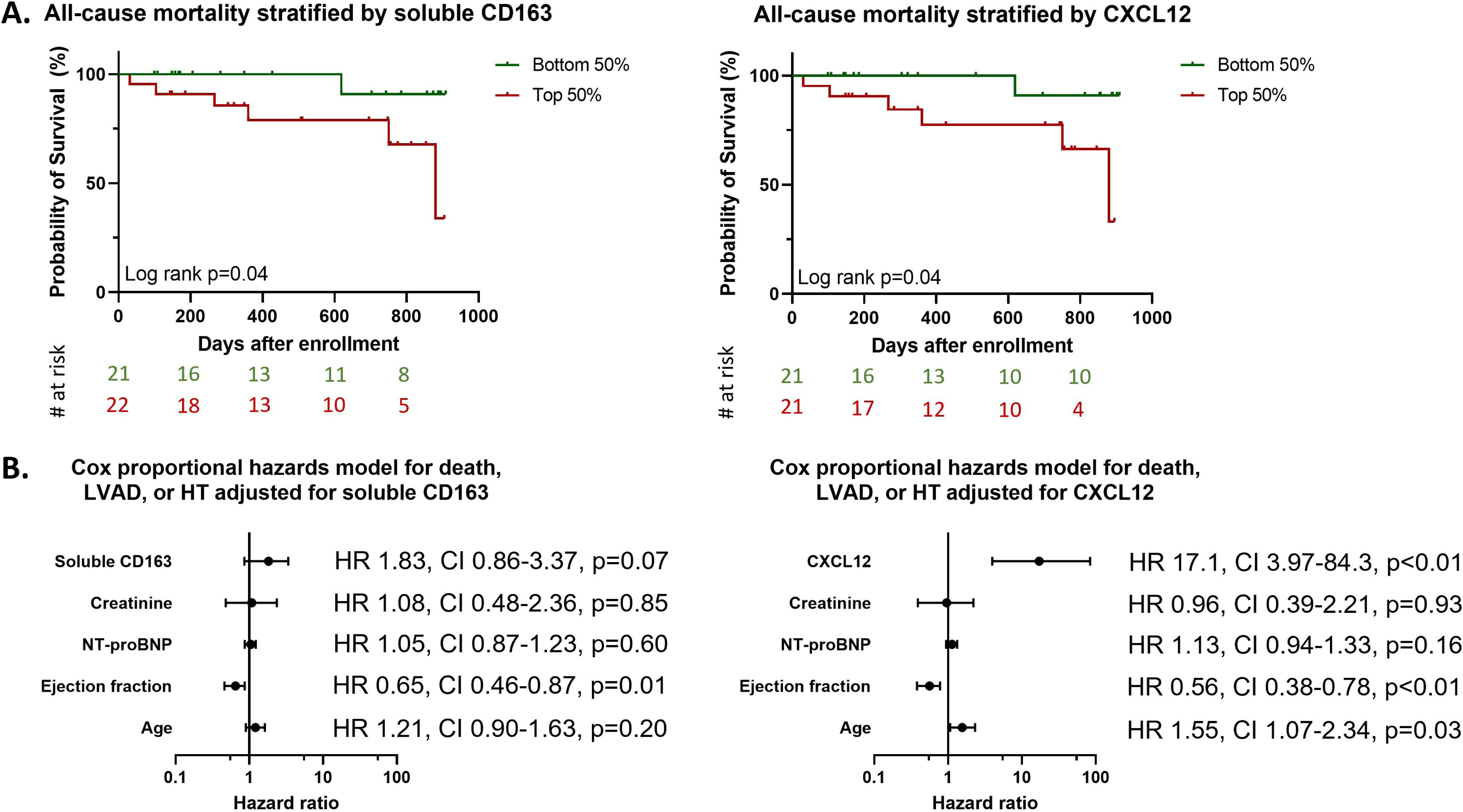
Soluble CD163 and CXCL12 are associated with worse outcomes. **(A)** Kaplan-Meier curves for all-cause mortality, stratified by median value of soluble CD163 and CXCL12. **(B)** Cox proportional hazards model for death, LVAD, or transplant adjusted for soluble CD163/CXCL12, creatinine, NT-proBNP, ejection fraction, and age. The Hazard ratios, 95% confidence intervals, and p-values are shown.

### Validation of sCD163 and CXCL12 in an independent HF cohort

To validate our findings in an external cohort, we analyzed data obtained from 172 patients of the Washington University HF Registry who underwent SOMAscan proteomic assessment. These were ambulatory HF patients who were less sick compared to those in our derivation cohort. Yet, the KM curves stratified by quartiles in this larger, healthier population showed that patients with the highest sCD163 and CXCL12 relative fluorescence unit appear to have decreased HT-free survival **(Figure 5A)**. In multivariable Cox proportional hazards model, sCD163 and CXCL12 were independently associated with death or need for HT **(Figure 5B)**. VEGF-A, on the other hand, did not predict clinical outcomes in this validation cohort **(Supplemental Figure 2)**.

**Figure 5.**
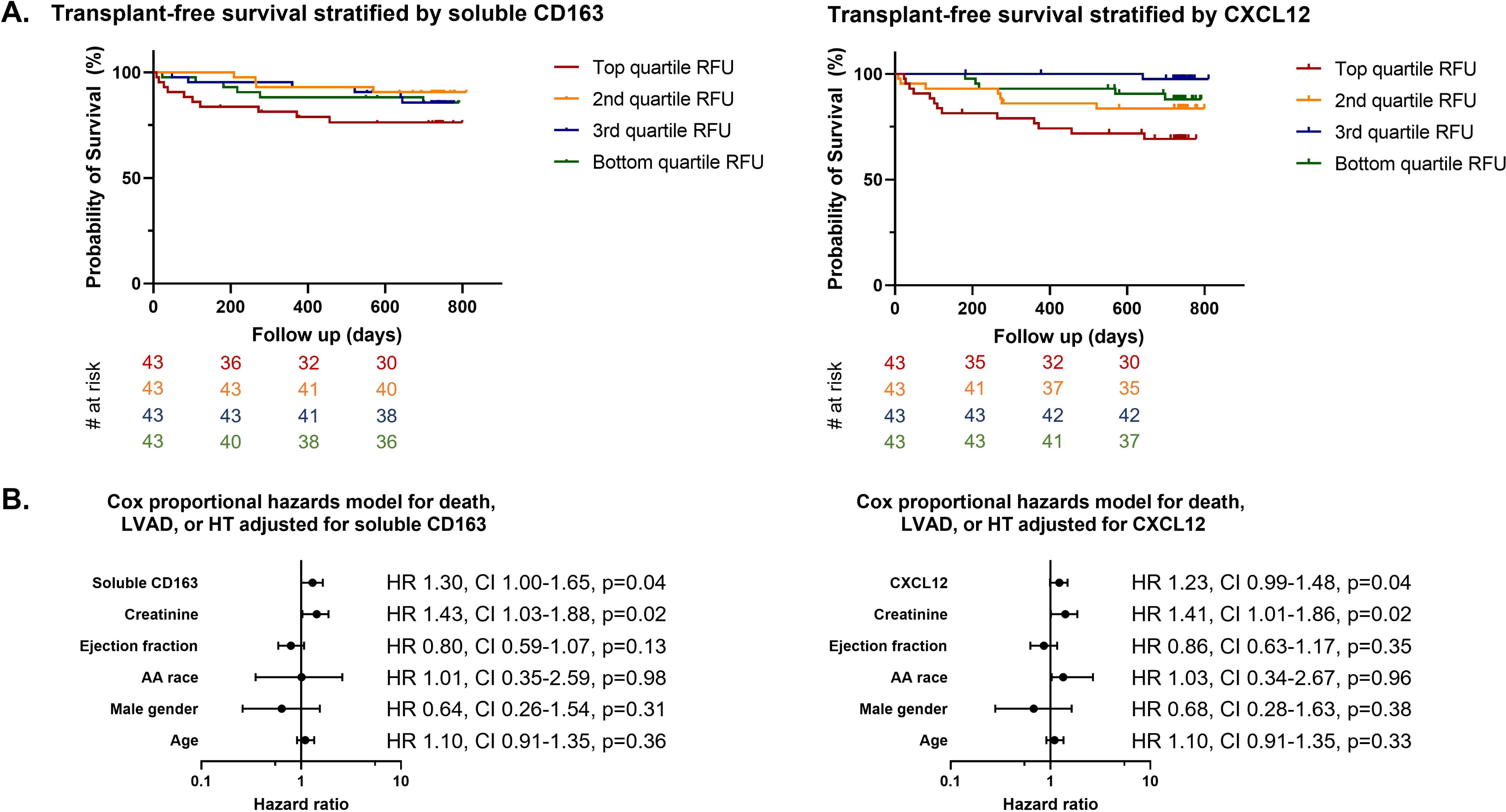
Validation of soluble CD163 and CXCL12 as markers of worse outcomes in patients with heart failure. **(A)** Kaplan-Meier curves for transplant-free survival in the Washington University Heart Failure Registry, stratified by quartiles of soluble CD163 and CXCL12. **(B)** Cox proportional hazards model for transplant-free survival adjusted for soluble CD163/CXCL12, creatinine, ejection fraction, race, gender, and age. The Hazard ratios, 95% confidence intervals, and p-values are shown. RFU=relative fluorescence unit, AA=African-American.

### Correlation of biomarkers to clinical and hemodynamic variables

Based on the relationship between these markers and mortality, we performed correlational analysis of sCD163 and CXCL12 with hemodynamic, echocardiographic, and laboratory values **(Supplemental table 1).** Neither factor correlated with cardiac index, LVEF, LV end diastolic diameter, or NT-proBNP. The strongest association was seen between sCD163 and bilirubin (r=0.63, p<0.01) and CXCL12 with RAP and RAP/PCWP ratio (r=0.61 and 0.57 respectively, p<0.01).

### sCD163 and CXCL12 are expressed by KC

As the liver is impacted by elevated CVP produced by RHF, we hypothesized that these circulating factors may reflect the hepatic response to HF **(Figure 6, central illustration)**. Using recently published scRNAseq data from human liver tissue, we confirmed that amongst cells in the liver, CD163 is uniquely expressed by KC. In addition, we discovered that CXCL12 was also highly expressed by KC and to a lesser extent by stromal cells **(Figure 6A)**. Based on these observations, we hypothesized that liver congestion may impact KC biology. To evaluate this concept, we stained human liver tissue from normal patients and those with CH for the KC marker CD163 and CXCL12. Compared to normal controls, patients with HF and CH had a higher density and increased quantity of CD163+ KC. Moreover, CD163+ KC appeared larger and often formed aggregates in CH livers, consistent with an altered activation state **(Figure 6B-C)**. Moreover, to confirm the expression of CXCL12 protein in human KC, we performed immunofluorescence and observed colocalization of CXCL12 with CD68+ KC in the liver parenchyma (**Figure 6D**). Taken together, these findings support KC as a potential source of CD163 and CXCL12 within the liver (**Figure 6E**).

**Figure 6.**
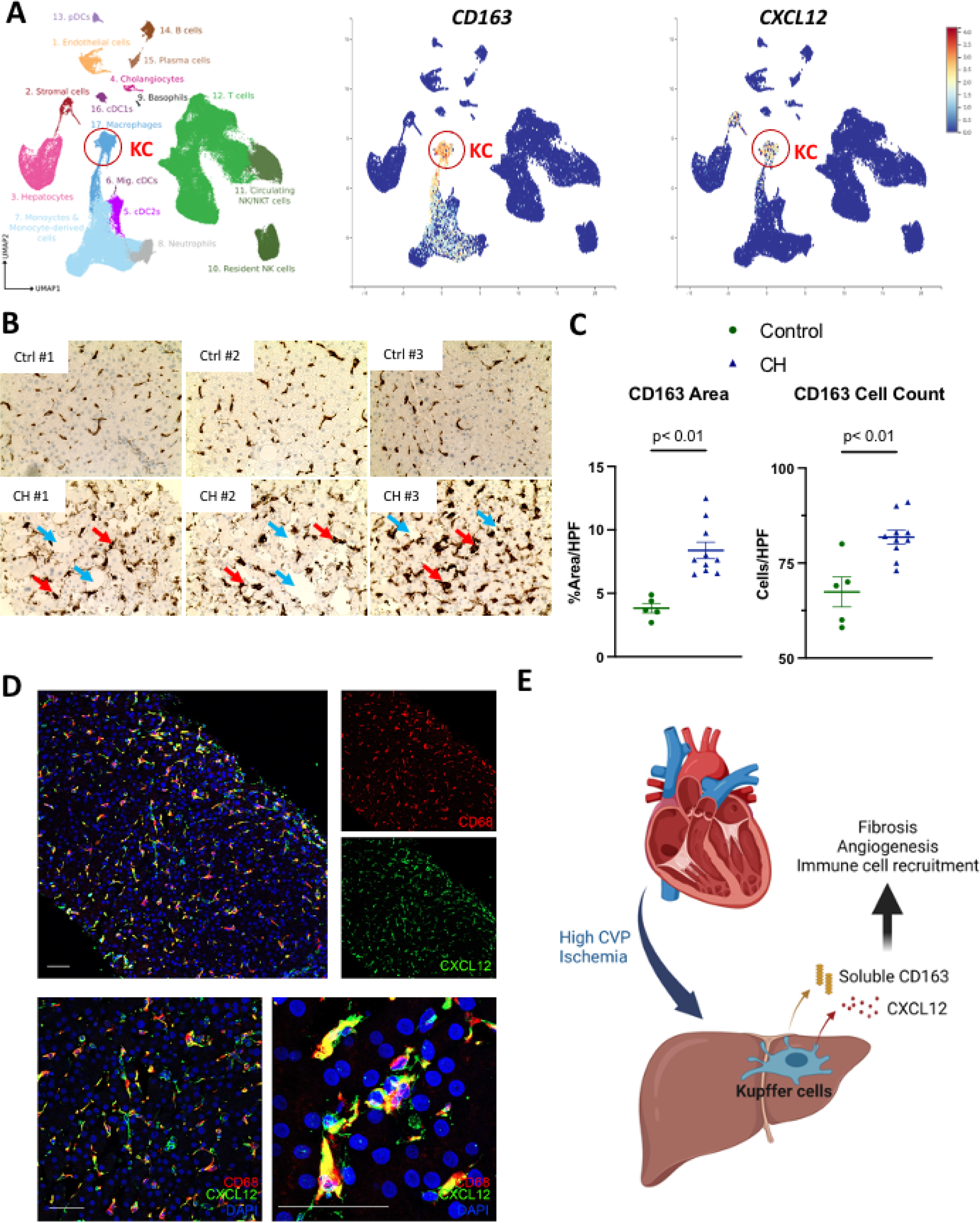
Kupffer cells (KC) are impacted in congestive hepatopathy (CH). **(A)** t-SNE plot generated from single cell RNA sequencing of human liver demonstrating clusters according to cell population (left). KC are indicated by the red circle. t-SNE dot plot expression of CD163 and CXCL12 for live liver cells, highlighting their preferential expression in KC (right). **(B)** Representative immunohistochemistry of CD163 on liver tissue from controls (top) and those with CH (bottom). Red and blue arrows denote KC and dilated sinusoids characteristic of CH, respectively. **(C)** Quantification of CD163 cell number and area in controls versus CH patients. **(D)** Immunofluorescence imaging of CXCL12 and CD68 in liver tissue from patients with CH demonstrating significant colocalization. Low power (upper panels) and high power (lower panels) are shown. Scale bars are 50 microns. **(E)** Schematic representation of right heart failure impacting the release of CD163 and CXCL12 from liver KC. HPF=high powered field.

## DISCUSSION

In this study, we evaluated the circulating inflammatory milieu in patients with RHF and found an elevation of several cytokines and growth factors. In particular, we identified sCD163 and CXCL12 as biomarkers with prognostication value in two separate cohorts. These data indicate that RHF is associated with a unique profile of circulating inflammatory factors that independently predict adverse outcomes in HF patients.

Previous studies have shown that elevated levels of circulating proinflammatory cytokines can be found in patients with HF.^9, 19–22^ However, most of these studies have focused on a small number of classic pro-inflammatory cytokines such as TNFα and IL-6.^23, 24^ Even less is known about the inflammatory milieu in RHF as this patient population has not been specifically studied. To gain insight into the secretome of RHF, we measured a total of 46 factors that represented a spectrum of cytokines, chemokines, and growth factors in three cohort of patients: 1) those without HF, 2) those with predominant left-sided HF, and 3) those with RHF who met prespecified criteria of right ventricular (RV) impairment. The majority of patients in this study had advanced HF, as evidenced by 25% LVAD/HT-free survival by 2 years of follow up. In line with other studies, we found that cytokines previously associated with HF such as TNFα and CXCL10 were elevated in both our HF and RHF cohorts.^20, 25^ However, we identified two factors, sCD163 and CXCL12, that have not been previously reported in HF literature that were elevated in RHF and were predictive of LVAD/HT-free survival in two separate HF populations. These observations support the notion that the syndrome of RHF is associated with alterations in immune activation state.

CD163 is a scavenger receptor for hemoglobin-haptoglobin complexes that is expressed by resident macrophages and can be cleaved and released into circulation as a soluble molecule in response to inflammatory cues.^26, 27^ KC are resident macrophages of the liver and represent ∼80% of the total macrophage pool in the human body.^28^ Localized within the vascular lumen of the hepatic sinusoids, KC are primed to release factors into systemic circulation. We used scRNAseq data and immunohistochemistry to confirm that CD163 is uniquely expressed by KC within the liver. In addition, KC density and size were increased in biopsy specimen from patients with CH compared to normal controls. In other chronic liver diseases such as cirrhosis and portal hypertension, sCD163 has also been found to correlate with prognosis.^29, 30^ Furthermore, we noted a correlation between sCD163 and serum bilirubin level, supporting the biological plausibility that sCD163 is of liver-origin. Taken together, these observations suggest that higher sCD163 in RHF may result from cleavage of CD163 from the surface of KC, possibly in response to inflammatory cues from the gut and/or due to mechanical stress from volume overload.

Our analysis also highlighted another unique factor that was elevated in patients with RHF, the chemokine CXCL12, also known as stromal derived factor-1 (SDF-1). CXCL12 has been described to play a role in diverse biologic processes including vascular development, bone marrow stem cell retention, and fibrosis.^31, 32^ In addition, CXCL12 has been shown to alter cardiac remodeling after myocardial infarction and predict incident HF in patients from the Framingham study.^33, 34^ Relevant to our findings in RHF, CXCL12 levels are also elevated in patients with pulmonary arterial hypertension and inhibition of this chemokine improves pulmonary hemodynamics and RV remodeling in animal models.^35, 36^ Furthermore, liver injury induces the expression of CXCL12, which can regulate hepatic fibrosis.^37, 38^ Using human liver scRNAseq data and immunofluorescence, we show that even under baseline conditions, KC are the primary hepatic cell population expressing this chemokine, providing further evidence linking CH and KC dysfunction. The functional role of CXCL12 in liver pathology and HF progression remains to be elucidated.

Our study has several limitations. First, we could not ethically perform an invasive procedure on healthy volunteers. Therefore, patients without HF were used as controls. Many of these patients underwent RHC as part of lung transplant evaluation and their underlying pulmonary pathology (i.e. idiopathic pulmonary fibrosis) could have attenuated differences between controls and HF patients. However, this would have been anticipated to decrease any observed differences in inflammatory profile between our control and HF cohorts. Second, patients with RHF may represent a subset of individuals with more severe disease. Arguing against this notion was that both HF and RHF patients had similar indications for RHC, similar laboratory findings and cardiac indices, and similar LVAD/HT-free survival. Thus, it is less likely that these observations simply represent a “sicker” patient population, but rather were a signature of RHF. Lastly, this is a small, single center study comprised mostly of patients with advanced HF and therefore our results cannot be generalized to all HF patients. Nevertheless, we validated our results in an ambulatory HF cohort and found that sCD163 and CXCL12 independently predicted HT-free survival in this healthier population.

## CONCLUSION

Patients with RHF have a distinct systemic proinflammatory profile compared to HF and normal controls. sCD163 and CXCL12 predicted mortality and LVAD/HT-free survival in both an advanced and ambulatory HF population. Future studies should focus on identifying the source of these factors and the systemic consequences of elevated sCD163 and CXCL12 in HF.

## Data Availability

All data is available for open access from the corresponding author.

## ACKNOWLEDGMENTS

We would like to thank the Immunomonitoring and the Anatomic and Molecular Pathology Core Labs at Washington University for performing the multiplex assays and immunohistochemistry.

## SUPPLEMENTAL MATERIALS

Tables S1

Figures S1-S2

## SOURCES OF FUNDING

BQY is funded by T32HL007081-41. This study was supported by American College of Cardiology, Missouri Chapter Research Grant and the Washington University Institute of Clinical and Translational Sciences grant UL1TR002345 from the National Center for Advancing Translational Sciences (NCATS) of the National Institutes of Health (NIH).

## DISCLOSURES

All authors report that there are no relevant disclosures.

## Notes

### Competing Interest Statement

The authors have declared no competing interest.

### Funding Statement

NIH T32HL007081-41 and UL1TR002345 from the National Center for Advancing Translational Sciences (NCATS) of the National Institutes of Health (NIH).

### Author Declarations

Washington University School of Medicine IRB

